# Clinical and intestinal histopathological findings in SARS-CoV-2/COVID-19 patients with hematochezia

**DOI:** 10.1101/2020.07.29.20164558

**Authors:** Margaret Cho, Weiguo Liu, Sophie Balzora, Yvelisse Suarez, Deepthi Hoskoppal, Neil Theise, Wenqing Cao, Suparna A. Sarkar

## Abstract

Gastrointestinal (GI) symptoms of SARS-CoV2/COVID-19 in the form of anorexia, nausea, vomiting, abdominal pain and diarrhea are usually preceeded by respiratory manifestations and are associated with a poor prognosis. Hematochezia is an uncommon clinical presentation of COVID-19 disease and we hypothesize that older patients with significant comorbidites (obesity and cardiovascular) and prolonged hospitalization are suspectible to ischemic injury to the bowel.

We reviewed the clinical course, key laboratory data including acute phase reactants, drug/medication history in two elderly male patients admitted for COVID-19 respiratory failure. Both patients had a complicated clinical course and suffered from hematochezia and acute blood loss anemia requiring blood transfusion around day 40 of their hospitalization. Colonoscopic impressions were correlated with the histopathological findings in the colonic biopies and changes compatible with ischemia to nonspecific acute inflammation, edema and increased eosinophils in the lamina propria were noted. Both patients were on anticoagulants, multiple antibiotics and antifungal agents due to respiratory infections at the time of lower GI bleeding. Hematochezia resolved spontaneously with supportive care. Both patients eventually recovered and were discharged.

Elderly patients with significant comorbid conditions are uniquely at risk for ischemic injury to the bowel. Hypoxic conditions due to COVID-19 pneumonia and respiratory failure, compounded by preexisting cardiovascular complications, and/or cytokine storm orchestrated by the viral infection leading to alteration in coagulation profile and/or drug/medication injury can be difficult to distinguish in these critically ill patients. Presentation of hematochezia may further increase the mortality and morbidity of COVID-19 patients, and prompt consultation and management by gastroenterology is therefore warranted.

## Introduction

In January 2020, molecular based studies identified the infectious agent SARS-CoV-2/COVID-19 (novel coronavirus 2019) from a cluster of cases of pneumonia reported in December from Wuhan, Hubei Province, China [1, 2] which has caused a worldwide pandemic. The viral infection typically manifests initially by cough and fever that rapidly escalates to pneumonia and respiratory failure in some patients [3] especially in the older population (>75 years old), leading to increased morbidity and mortality [4]. Gastrointestinal symptoms usually present as anorexia, nausea, vomiting, abdominal pain, anosmia and hypogeusia, and diarrhea and are associated with poor prognosis [5]. The possibility of fecal-oral transmission is also supported by the fact that virus can be identified in the stool and rectal swabs of infected patients even with a negative nasopharyngeal swab test [6, 7]. It has also been shown that SARS-CoV-2 uses angiotensin-converting enzyme (ACE2) as a viral receptor for the entry process [8]. By using quantitative polymerase chain reaction, researchers have shown that high levels of ACE2 mRNA expression can be detected in the gastrointestinal tract especially the small bowel and the colon providing the likely route for viral infection [8]. In this report, we present the clinical course, colonoscopy findings, corresponding histopathological changes, and pertinent laboratory findings in two patients with SARS-CoV-2/COVID-19 pneumonia, complicated by an uncommon presentation of hematochezia during their prolonged hospitalization.

## Case Report/Case Presentation

### Clinical history and hospital course for case 1

A 67-year old man with a past medical history of obesity (108.9kg, Body Mass Index 38.45 kg/m^2^), diabetes mellitus type 2, hypertension, hyperlipidemia, and left bundle branch block presented to the emergency department with fever and chills for 8 days and cough with shortness of breath for 3 days. The patient received hydroxychloroquine and azithromycin from his primary care provider for presumed COVID-19 infection but discontinued by the patient after a few days. On admission, nasopharyngeal swab was positive for SARS-CoV-2 virus. COVID-19 pneumonia was complicated by acute hypoxic respiratory failure requiring intubation. His nasal swab was also positive for methicillin-susceptible staphylococcus aureus. The patient’s hospital course was further complicated by a non-ST-elevation myocardial infarction, atrial fibrillation with rapid ventricular rate, diabetes insipidus, toxic encephalopathy, acute kidney injury, and right upper and lower extremity weakness. He also developed acute occlusive deep vein thrombosis of his right peroneal vein for which he received apixaban. His medications included azithromycin, hydroxychloroquine, ceftriaxone, fluconazole, vancomycin, piperacillin/tazobactam, and cefazolin.

On day 33 of hospitalization, the patient passed frank red blood per rectum coupled with hemodynamic instability requiring blood transfusion. Colonoscopy was performed to identify the source of bleeding. At the time of gastrointestinal (GI) bleeding, he was on pantoprazole, albuterol, aspirin (low dose) fluconazole, furosemide, oxycodone, and piperacillin-tazobactam. Stool tests was negative for pathogens. Anticoagulation was stopped temporarily in the setting of active bleeding.

### Colonoscopy findings

Notable findings included a few scattered non-bleeding erosions and shallow ulcerations in the transverse colon, hepatic flexure, and ascending colon (shown in Figure 1a). Biopsies of the ulcerated mucosa and of normal mucosa were obtained. The terminal ileum appeared normal. The source of hematochezia was considered to be from the small scattered proximal colonic erosions and shallow ulcers, exacerbated by patient’s use of apixaban.

**Figure 1.**
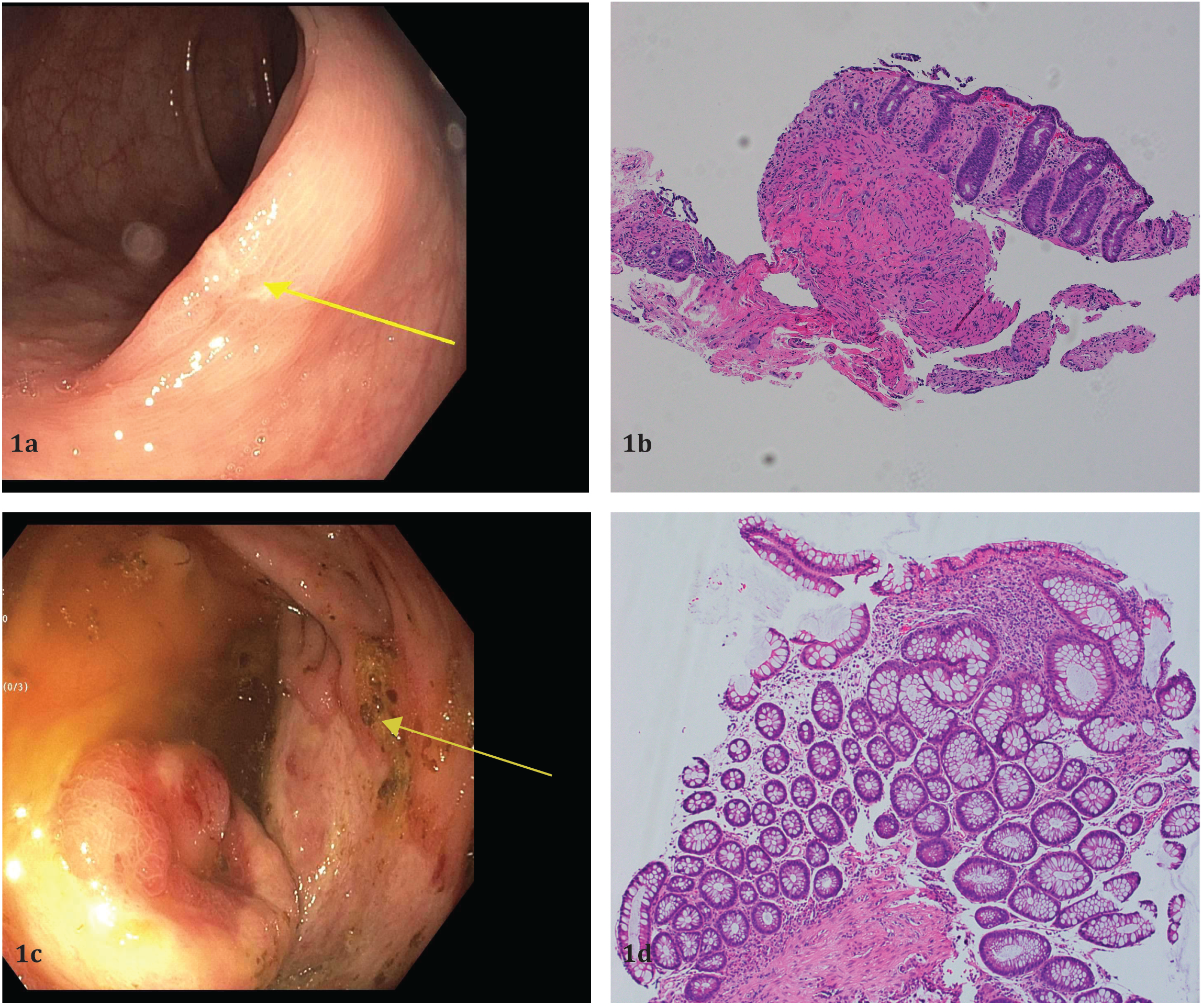
Colonoscopic findings **(a, c)** and histopathology of biopsies **(b, d)** counterstained with haematoxylin and eosin (x 100), **(a)** Case 1, transverse colon with erosion (yellow arrow), **(b)** Case 1, colonic mucosa with erosion, withered crypts, and nonspecific acute inflammation, consistent with ischemic pattern of injury, **(c)** Case 2, ileocecal valve with erosions (yellow arrow), **(d)** Ileocolic mucosa with edema, mildly active nonspecific inflammation and focally increased eosinophils in the lamina propria.

### Histopathologic findings

The colonic mucosa featured an erosion, withered crypts, and nonspecific acute inflammation (shown in Figure 1b). The overall findings were consistent with an ischemic pattern of injury. Alternate possibilities of infection and drug/medication mediated mucosal injury were raised. No specific features of chronicity were identified. No viral cytopathic changes or microthrombi were seen. An immunohistochemical stain for cytomegalovirus was negative.

### Outcome

On day 18, the patient had a tracheostomy placement and was decannulated on day 35. GI bleeding resolved spontaneously, and anticoagulation was resumed. On day 49 he was discharged home with rehabilitation.

### Clinical history and hospital course for case 2

A 68-year old man with a past medical history of tobacco use, obesity (81.6 kg Body weight, 29 kg/m^2^ BMI), diabetes mellitus 2, hypertension, hyperlipidemia, and coronary artery disease was brought to the emergency department for cough, fever, loose stools and altered mental status for five days. On admission, nasopharyngeal swab was positive for SARS-CoV-2 virus by PCR. COVID-19 pneumonia was complicated by acute hypoxic respiratory failure requiring intubation. His clinical course was complicated by sepsis, acute kidney injury requiring peritoneal dialysis, ileus, toxic metabolic encephalopathy, and candidemia (candida tropicalis and candida glabrata). On day 40 of his hospitalization, intermittent hematochezia requiring blood transfusion were noted. A subset of his critical laboratory parameters is highlighted in Table 2. CT abdomen pelvis showed the bowel was of normal caliber with no evidence of obstruction. Colonoscopy was performed to identify the source of bleeding. At the time of the bleeding, the patient was being treated with fluconazole, meropenem, micafungin, intravenous vancomycin, and ampicilin-sulbactum.

### Colonoscopy findings

Patchy, deeply ulcerated mucosa with significant edema and decreased vascular pattern was present in the cecum with involvement of the ileocecal valve (shown in Figure 1c). There was diffuse oozing of the mucosa and the endoscopic appearance of the mucosa was clinically consistent with ischemic colitis. The ileocecal valve showed significant edema and ulceration. Biopsies were taken with cold forceps for histologic evaluation. Normal mucosa was found in the remaining portions of the colon.

### Histopathological findings

The cecal mucosa showed focal edema, mildly active nonspecific inflammation and focal increased eosinophils in the lamina propria. There was no definitive evidence of ischemia (shown in Figure 1d).

### Clinical outcome

Gastrointestinal bleeding resolved spontaneously. Patient was decannulated on day 54 and admitted to the rehabilitation unit.

### Discussion/Conclusion

Gastrointestinal manifestations of COVID-19 are seen more often than originally suspected in COVID-19 patients [9-11].The cases presented are male patients above 65 years of age with complicated and prolonged hospitalizations. Although both patients presented with severe COVID-19 pneumonia resulting in hypoxic and respiratory failure on arrival, only one presented with gastrointestinal symptoms in the form of diarrhea. Both patients were obese and with cardiac comorbidities that are known to increase the morbidity and mortality in such cases [12]. The hospital courses for both patients were remarkable for lower GI bleeding in the latter part (>40 days) of hospitalization. Colonoscopies were remarkable for scattered erosions and ulceration clinically compatible with ischemia. The pathologic findings of the GI tract showed nonspecific acute inflammation and ischemic pattern of injury; however, drugs/medication, infections could not be completely ruled out. Both patients were on multiple medication, anticoagulants and antibiotics and antifungal agents at the time of hematochezia. Lower GI bleeding is an uncommon manifestation of COVID-19 and has been recently reported as a presenting symptom in an elderly male [13]. It is unclear if the GI symptoms of COVID-19 infection are caused by thrombosis due to altered coagulation profile (shown in Table 1 and 2) or as a consequence of tissue damage secondary to an immune response, rather than a direct interaction of these epithelial cells with the virus [14]. Additionally, an abnormal coagulation profile featuring significantly elevated D-dimer, fibrin degradation product levels, increased prothrombin time and activated partial thromboplastin time have also been reported in COVID-19 patients [15] and was also seen in our patients.

**Table 1.** Pertinent laboratory 1 findings from Case 1.

|  | Admission (Day 1) | Peak | Discharge (Day 49) |
| --- | --- | --- | --- |
| Cobas SARS-CoV-2 real time RT-PCR | Detected | N/A | Not detected (retested on day 40 and 42) |
| Hemoglobin | 13.9 g/dL | 6.7 g/dL (day 40-41)<br>Intermittent Rectal Bleeding (day 33) | 9.0 g/dL |
| WBC | 9.7 $10^3/\mu\text{L}$ | 28.4 $10^3/\mu\text{L}$ | 5.4 $10^3/\mu\text{L}$ |
| D-Dimer | 284 ng/ml DDU | 3414 ng/ml DDU (day 24) | 375 ng/ml DDU |
| Lactate dehydrogenase | 398 U/L | 463 U/L (day 11) | 264 U/L |
| Procalcitonin | 0.13 ng/ml | 0.75 ng/ml (day 35) | 0.08 ng/ml |
| C-Reactive Protein | 154.4 mg/L | 163.6 mg/L (day 2) | 23.5 mg/L |
| AST | 56 U/L | 461 U/L (day 18) | 108 U/L |
| ALT | 49 U/L | 629 U/L (day 18) | 198 U/L |
| Alkaline<br>Phosphatase | 44 U/L | 1521 U/L (day 40) | 1370 U/L |
| Bilirubin Direct | 0.3mg/dL | 0.7mg/dL (day 18) | 0.4mg/dL |
| Creatinine | 0.82mg/dL | 5.15mg/dl (day<br>13) | 0.62mg/dL |
| Blood Glucose | 101mg/dL | 367mg/dL (day<br>10) | 175mg/dL |

**Table 2.** Pertinent laboratory 1 findings from Case 2.

|  | Admission (day 1) | Peak | Discharge (day 49) |
| --- | --- | --- | --- |
| Cobas SARS-CoV-2 real time RT-PCR | Detected<br>Detected again day 43 | N/A | Not detected (day 44) |
| Hemoglobin | 12.8 g/dL | 6.83g/dL (day 41<br>*Colonoscopy (day 40) | 8.04 g/ dL (day 53) |
| WBC | 5.5 10<sup>3</sup>/μL | 26.2 10<sup>3</sup>/μL (day12) (day 12) | 9.1 10<sup>3</sup>/μL (day 53) |
| D-Dimer | 456 ng/ml DDU | 4536 ng/ml DDU (day 12) | 555 ng/ml DDU (day 53) |
| Lactate dehydrogenase | 273 U/L | 1134 U/L (day 11) | 336 U/L |
| Procalcitonin | 0.14 ng/mL | 7.59 ng/mL (day 14) | 0.45 ng/dL (day 34) |
| C-Reactive Protein | 150.3 mg/L | 382.4 mg/L (day 3) | 50.8 mg/L (day 50) |
| AST | 25U/L | 420 U/L (day<br>12) day12) | 40 U/L (day 51) |
| ALT | 49 U/L | 629 U/L (day<br>1812) | 198 U/L (day 51) |
| Alkaline<br>Phosphatase | 47 U/L | 695 U/L (day<br>47) | 570U/L (day 51) |
| Bilirubin Direct | 0.3 mg/dL | 0.4 mg/dL<br>(day 18) | 0.2 mg/dL |
| Creatinine | 1.06 mg/dL | 7.66 mg/dl<br>(day 13) | 1.65 mg/dL |
| Blood Glucose | 181 mg/dL | 414 mg/dL<br>(day 12) | 160 mg/dL |

To our knowledge, this is the first histopathological and clinical correlation from colonic biopsies of two patients with severe COVID-19 pneumonia complicated by hematochezia during the later phase of their prolonged hospitalizations. Potential etiologies for the colonoscopic and histopathological findings include thromoembolic events specifically microthrobi which may be evident in resection and autopsy specimen rather than biopsy samples, or related to damage caused directly by the viral cytopathic effect or related to the cytokine storm, ischemia/hypotension injury or a drug-induced injury. Careful review of medications, management of coagulation profile and consultation with gastroenterology is warranted to improve morbidity and mortality outcomes.

## Data Availability

All data for the case report are available and uploaded.

## Statements

## Acknowledgement

The autors would like to thank the Department of Pathology and the Division of Gastroenterology and Hepatology, NYU Langone Health for their support and assitance.

## Statement of Ethics

Every precaution has been taken to protect the privacy of research subjects and the confidentiality of their personal information. The study protocol was approved by the institute’s committee on human research. All patients in this report are identified by numbers.

## Disclosure Statement

The authors have no conflicts of interest to declare.

## Funding Sources

No funding source to report

## Author Contributions

MC, WL interpreted histopathology, SB performed colonoscopies on both cases and provided interpretation of clinical data, DH, WL, SAS contributed to acquisition of micrographs, MC, WL, SB, DH, YS, NT, WC interpreted data, edited manuscript, provided intellectual input. SS planned, reviewed chart, wrote the IRB and manuscript.

## Notes

### Competing Interest Statement

The authors have declared no competing interest.

### Funding Statement

The authors have no conflicts of interest to declare
No third party payments were received by for any aspect of the work by the authors and the institution.

### Author Declarations

Approved by IRB protocol from NYU Grossman School Of Medicine.

